# Income and Race-Ethnicity Disparities in Medical Care Utilization and Expenditures in the United States, 2017-2019

**DOI:** 10.1101/2022.05.31.22275747

**Authors:** Harry Wetzler, Herbert Cobb

## Abstract

**Background:** The COVID-19 pandemic has focused attention on race and income disparities in SARS-CoV-2 mortality and morbidity. Much less attention has been paid to other socioeconomic factors including income.

**Objective:** The goal of this study was to compare disparities in medical care utilization and related expenditures associated with income to those associated with race and ethnicity in the US for those aged 0 to 64 for four categories of medical services: hospital, emergency room, ambulatory care, and prescription medications.

**Methods:** We used Medical Expenditure Panel Survey data for years 2017 through 2019. For each of the four medical services, there were three measures. First was the percentage of those aged 0-64 with or without utilization and expenditures. Due to statistical issues related to zero values for utilization and expenditures, the second and third measures were average utilization and expenditures only for those with both utilization and expenditures. Disparities by income and race-ethnicity were measured by calculating the percent difference between the group with the lowest utilization or expenditures and the group with the highest utilization or expenditures.

**Results:** For 9 of the 12 separate differences the income differences exceed the corresponding race-ethnicity difference and the income differences are generally much greater in magnitude. Within the income comparisons, those on Medicaid had the greatest utilization in 7 of the 8 comparisons. The High Income group had greatest expenditures for 3 of the 4 medical services. Non-Hispanic Whites had the greatest utilization and expenditures for 9 of the 12 measures and Hispanics had the least utilization and expenditures for 9 of the 12 measures.

**Conclusions:** These results indicate that income inequalities are more strongly associated with medical care utilization and expenditures than race-ethnicity among those aged 0-64. Although more research should focus on income related health disparities in the United States, it is time to recognize that sound health policy must include reducing socioeconomic inequalities, especially those related to income.

## INTRODUCTION

Health disparities manifest themselves in diverse ways from variations in mortality and morbidity to differences in medical care utilization and expenditures. Recently, decreases in life expectancy that differ markedly by race and ethnicity occurred during the COVID-19 pandemic.^1^ Between 2019 and 2020, life expectancy decreased by 3.0 years for the Hispanic population (81.8 to 78.8), 2.9 years for the non-Hispanic black population (74.7 to 71.8), and 1.2 years for the non-Hispanic white population (78.8 to 77.6). Dieleman and colleagues found that in 2016 US health care spending varied by race and ethnicity across different types of care even after adjusting for age and health conditions.^2^ For instance, White individuals received an estimated 15% more spending than the all-population mean for ambulatory care. Non-Hispanic Black individuals received an estimated 26% less spending on ambulatory care but 19% more on inpatient and 12% more on emergency department care. Hispanic individuals received an estimated 33% less spending per person on ambulatory care. However, Dieleman, et al, did not include income in their analyses. Race, ethnicity, and relative income are correlated. According to Meyer and Sullivan, poverty rates in Blacks and Latinos are approximately twice those in Whites.^3^

Chetty, et al. found differences in mortality by income level.^4^ In the United States between 2001 and 2014, the gap in life expectancy between the richest 1% and poorest 1% of individuals was 14.6 years for men and 10.1 years for women. Between 2001 and 2014, life expectancy increased by 2.34 years for men and 2.91 years for women in the top 5% of the income distribution, but by only 0.32 years for men and 0.04 years for women in the bottom 5%.

Mahajan and colleagues emphasized differences in health status by race and ethnicity in the US between 1999 and 2018.^5^ However, in 2017-2018 the *highest* reported difference in self-reported health status by race-ethnicity was 7.95 percentage points whereas the *lowest* reported difference by income within race-ethnic groups was 8.5 percentage points. These findings suggest that income inequality has considerably more impact on health compared to race and ethnicity.^6^

The goal of this study was to compare disparities in medical care utilization and related expenditures associated with income to those associated with race and ethnicity in the US for those aged 0 to 64 for four categories of medical services: hospital, emergency room, ambulatory care, and prescription medications.

## MATERIALS AND METHODS

### Data Source

The Medical Expenditure Panel Survey (MEPS) is cosponsored by the Agency for Healthcare Research and Quality (AHRQ) and the National Center for Health Statistics (NCHS). MEPS is designed to provide nationally representative estimates of health care use, expenditures, sources of expenditure, and health insurance coverage for the U.S. civilian non-institutionalized population. Data on respondents’ demographic and socio-economic characteristics are also available. The MEPS Household Component (MEPS-HC) is a subsample of households participating in the previous year’s National Health Interview Survey (NHIS) conducted by the NCHS. The NHIS oversamples Asian, black, and Hispanic persons as well as policy relevant sub-groups such as low-income households.^7^ Although data for individual years are available from AHRQ, the University of Minnesota IPUMS MEPS site has collated data that were downloaded for this study.^8^ MEPS data are in the public domain.

MEPS is authorized under 42 U.S.C. 299a. Privacy is protected by the Privacy Act and Section 308(d) of the Public Health Service Act [42 U.S.C. 299c-3(c) and 42 U.S.C. 242m(d)], and respondents’ confidentiality is protected by Section 944(c). Individual identifiers are removed from the micro-data files before they are released to the public.

### Study sample

We used MEPS data for years 2017 through 2019. Most of the analyses were restricted to respondents aged 0 to 64 which removed potential confounding issues related to Medicare insurance. We calculated summary age and gender statistics for the groups described below.

### Variables

Income categories were based on total family income divided by the applicable federal poverty line (based on family size and composition including unmarried partners and foster children). The specific income categories were 0 to 1.99, 2 to 3.99, and 4 or greater times the poverty line. We refer to these income categories as Low Income, Middle Income, and High Income in this report. Respondents who had 10 months or more of Medicaid coverage in a calendar year were placed in a separate income category. Ten months of Medicaid coverage was found to be the amount that produced Medicaid sample percentages comparable to those in the 2017 and 2018 NHIS.^9^ For race-ethnicity, respondents were asked if the person’s main national origin or ancestry was Puerto Rican; Cuban; Mexican, Mexican American, or Chicano; other Latin American; or other Spanish. All persons whose main national origin or ancestry was reported in one of these Hispanic groups, regardless of racial background, were classified as Hispanic. All other persons were classified according to their reported race. For this analysis, the following classification by race and ethnicity was used: Hispanic, non-Hispanic Black (referred to as Black in this report), non-Hispanic White (referred to as White in this report), and non-Hispanic other (referred to as Other in this report). The Other category includes American Indian, Alaska Native, Asian or Pacific Islander, other race, and multiple races. Covariates included age, gender, and survey year. In addition, when income was the independent variable, race-ethnicity was included as a covariate, and vice versa.

### Outcomes

Outcomes consisted of utilization and expenditure data for four categories: hospitalization, emergency room, outpatient visits, and prescription medications. Hospital utilization consists of the total number of nights the person spent in the hospital during the current calendar year. Emergency room medical provider visits include encounters that took place primarily in emergency room settings and clinics and include visits to both physician and non-physician providers. Outpatient visits include visits to both physician and non-physician providers. Prescription medications are a count of all prescribed medications purchased during the year (including initial purchases and refills).

All expenditure data are the sum of direct expenditures made during the year including out-of-pocket expenditures and expenditures by private insurance, Medicaid, Medicare, and other sources in the respective categories. Hospital expenditure data include expenditures for hospital facility expenses (including direct hospital care, room and board, diagnostic and laboratory work, x-rays, and similar charges, as well as any physician services included in the hospital charge) and “separately billing doctor” expenses (including services provided to patients in hospital settings by providers like radiologists, anesthesiologists, and pathologists, whose charges are often not included in hospital bills). To provide context for the expenditure data, we estimated the proportion of all US medical spending attributed to those aged 0-64 and then the proportions in that age group attributed to the four medical services.

For each of the four medical services, there were three measures. First was the percentage of those aged 0-64 with or without utilization and expenditures. Due to statistical issues related to zero values for utilization and expenditures, the second and third measures were average utilization and expenditures only for those with both utilization and expenditures. Disparities by income and race-ethnicity were measured by calculating the percent difference between the group with the lowest utilization or expenditures and the group with the highest utilization or expenditures. We then used the expenditure proportions for each medical service to calculate weighted expenditure summaries for the income and race-ethnicity groups.

### Statistical analysis

We combined data for three years (2017-2019) to yield sample sizes large enough to generate reliable estimates for subpopulations. MEPS-HC samples in most years are not completely independent because households are drawn from the same sample geographic areas and many persons are in the sample for two consecutive years. Despite this lack of independence, it is valid to pool multiple years of MEPS-HC data and keep all observations in the analysis because each year of the MEPS-HC is designed to be nationally representative.^7^ The analytic weight variable was divided by three to provide estimates that reflect an annual average. This adjustment has no effect on point estimates because the weight variable is divided by a constant.

Substantial proportions of respondents have no medical expenditures or utilization during a given year (over 90% for hospitalization) and standard statistical models such as linear regression are inappropriate for the entire sample. Thus, for each study category we first modeled those with and without utilization and expenditures using logistic regression. For those with both utilization and expenditures in a category, we used a generalized linear model (GLM) with a Poisson distribution for utilization and a gamma distribution and log link for expenditures. Covariates were modelled using indicator (dummy) variables. Since this was an exploratory study, we report no standard errors or confidence intervals and performed no inferential statistical tests. The appropriate weights were used in the analyses. Statistical analyses were conducted using Stata version 16.0 and SPSS version 25. The Stata margins command was used to produce estimates of adjusted marginal means from predictions of the fitted models. The “asbalanced” option of the Stata margins command was used to specify that factor covariates be evaluated as though there were an equal number of observations in each level.

## RESULTS

There was a total of 90,853 respondents for all ages with 74,758 under age 65. In the US civilian non-institutionalized population, expenditures for hospitalization, emergency room, outpatient visits, and prescription medications accounted for 86.4% of total expenditures for those aged 0 to 64 and 65.1% of total expenditures for all ages. For those aged 0-64, hospital, emergency room, outpatient, and prescription medications accounted for 26.1%, 5.0%, 42.7%, and 26.2% of total expenditures respectively in that age group. Table 1 lists the mean ages and female percentages of each income and race-ethnicity group and their combinations.

**Table 1.** Age and gender distributions by income and race-ethnicity.

| <u>Income Groups</u> | <u>Mean Age</u> | <u>Percent Female</u> |
| --- | --- | --- |
| Medicaid | 22.5 | 54.8% |
| Low | 32.9 | 50.4% |
| Mid | 32.3 | 49.6% |
| High | 35.9 | 48.7% |

| <u>Race-Ethnic Groups</u> | <u>Mean Age</u> | <u>Percent Female</u> |
| --- | --- | --- |
| Hispanic | 28.1 | 49.3% |
| Black | 31.0 | 52.3% |
| White | 34.2 | 50.0% |
| Other | 29.9 | 51.2% |

| <u>Income-Race-Ethnic Groups</u> | <u>Mean Age</u> | <u>Percent Female</u> |
| --- | --- | --- |
| Medicaid-Hispanic | 18.6 | 52.6% |
| Medicaid-Black | 22.6 | 56.4% |
| Medicaid-White | 25.4 | 56.0% |
| Medicaid-Other | 23.4 | 54.0% |
| Low Income-Hispanic | 31.7 | 48.9% |
| Low Income-Black | 33.4 | 53.0% |
| Low Income-White | 34.0 | 50.3% |
| Low Income-Other | 30.6 | 50.0% |
| Middle Income-Hispanic | 30.3 | 46.8% |
| Middle Income-Black | 33.3 | 51.3% |
| Middle Income-White | 33.1 | 50.1% |
| Middle Income-Other | 30.6 | 50.6% |
| High Income-Hispanic | 33.5 | 48.5% |
| High Income-Black | 35.9 | 48.5% |
| High Income-White | 36.9 | 48.5% |
| High Income-Other | 32.3 | 50.6% |

Those in the Medicaid income category were over 10 years younger on average and the percentage of females was nearly five points higher. The average age of Hispanics was three years less than Blacks and six years less than Whites. Nearly half of Hispanics and Blacks were in the Medicaid or Low Income categories, but less than a quarter of Whites were in those categories.

### Hospital Utilization and Expenditures

Table 2 lists the adjusted percentages of the age 0 to 64 US population with any hospital use and the average nights and expenditures per person per year for those with hospital use and expenditures.

**Table 2.** Hospital Use and Expenditures by Income Group and Race-Ethnicity.

| **Income Category** | **Percentage with Use** | **Nights** | **Expenditures** |
| --- | --- | --- | --- |
| Medicaid | 6.6% | 7.5 | \$14,767 |
| Low | 5.0% | 5.7 | \$14,757 |
| Mid | 3.4% | 5.1 | \$22,522 |
| High | 2.5% | 4.3 | \$25,408 |

| **Race-Ethnicity** | **Percentage with Use** | **Nights** | **Expenditures** |
| --- | --- | --- | --- |
| Hispanic | 3.4% | 4.2 | \$18,847 |
| NH_Black | 4.5% | 6.9 | \$19,414 |
| NH_White | 5.2% | 5.8 | \$19,662 |
| NH_Other | 3.6% | 5.6 | \$17,332 |

Overall, 4.6% of the age 0-64 population had hospital nights and expenditures. Comparing highest and lowest overall hospital use, those in the Medicaid Income category used 159.5% more than those in the High Income category. Non-Hispanic Whites had 51.5% more than Hispanics. Among those with hospital utilization, hospital nights were 76.8% greater in the Medicaid group compared to the High Income group. This relationship is seen across all race-ethnicity groups with the Medicaid group having the highest number of hospital nights per person followed in income order of Low, Middle, and High (Figure 1). Blacks had the highest number of hospital nights per person followed by Whites, Other, and Hispanic. The average number of nights for Blacks was 66.4% greater than that for Hispanics. Detailed data are listed in the Supplemental Data.

**Figure 1.**
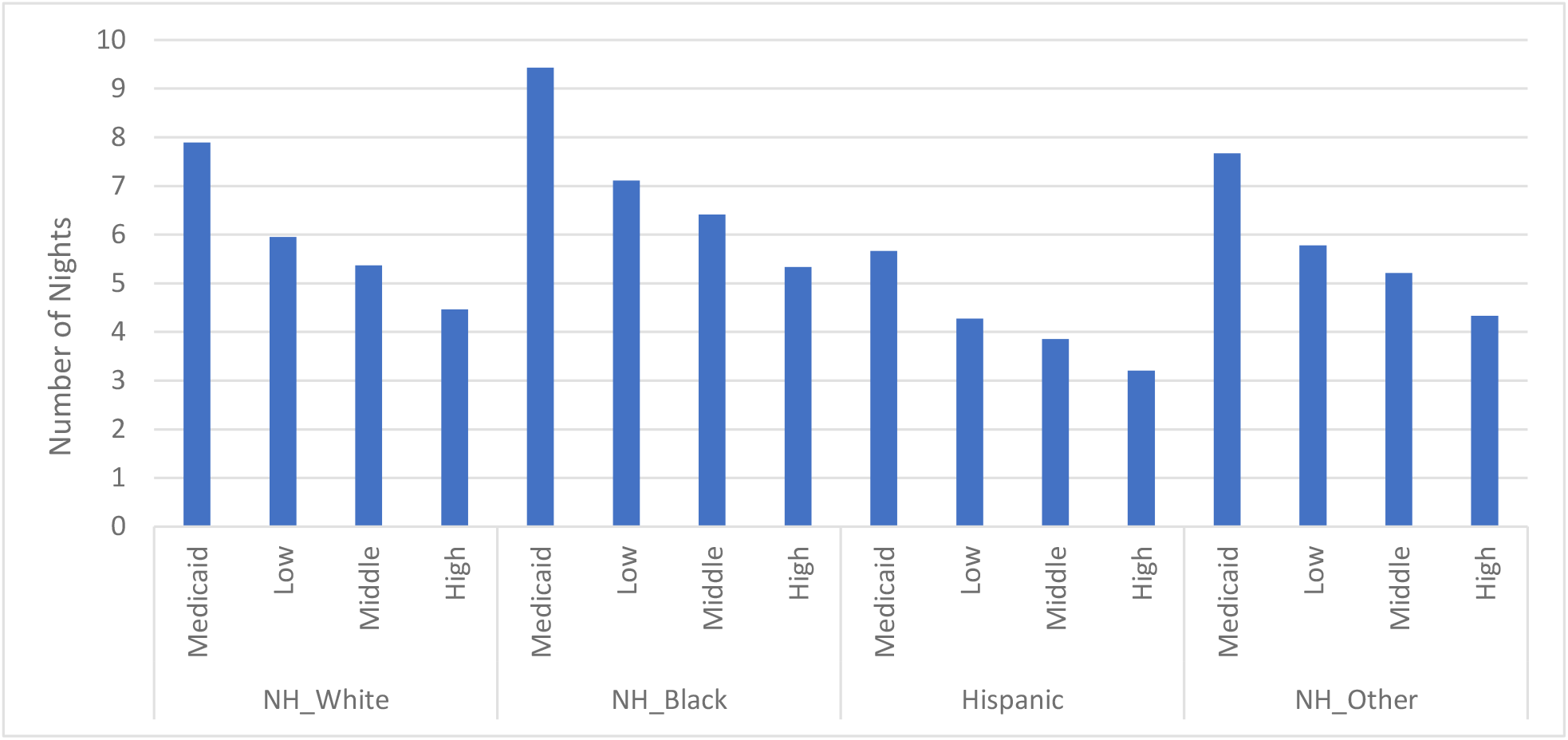
Hospital Nights per Person by Income and Race-Ethnicity. (for those with hospital nights and expenditures greater than zero).

Among those with any hospital utilization and expenditures, the expenditures per person for the High Income group were 72.2% higher than those for the Low Income group (note that hospital expenditures were almost the same for the Low Income and Medicaid groups). Race-ethnicity differences were similar across all income groups with the White group having the highest hospital expenditures per person followed by Black, Hispanic, and Other (Figure 2). The maximum race-ethnicity disparity was Whites compared to Others where the Whites’ expenditures were 13.4% higher.

**Figure 2:**
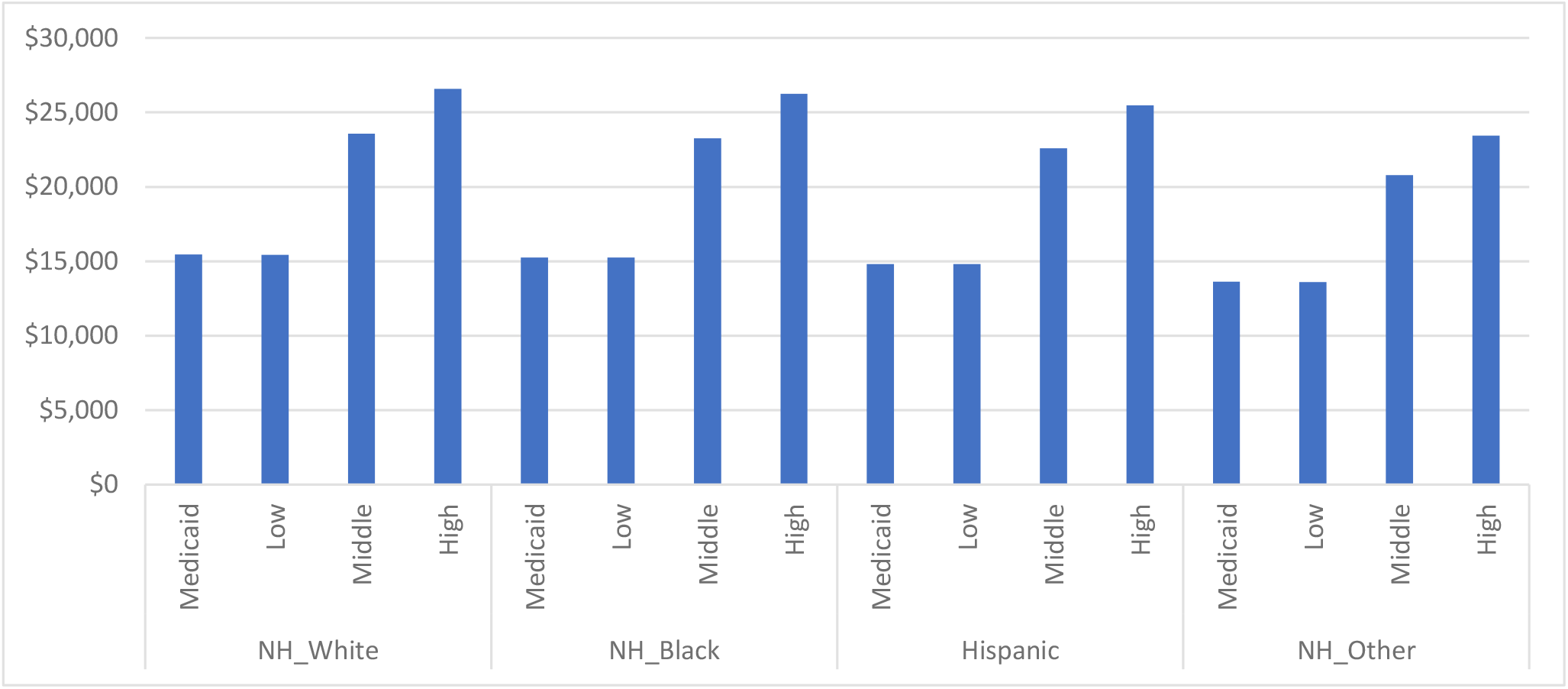
Hospital Expenditures per Person by Income and Race-Ethnicity. (for those with hospital nights and expenditures greater than zero).

### Emergency Room Visits and Expenditures

Table 3 lists the adjusted percentages of the age 0 to 64 population with any emergency room use and the average visits and expenditures per person per year for those with emergency room use and expenditures (note that the average visits by race-ethnicity are all 1.4, a coincidence due to rounding; the actual range is 1.356 to 1.417).

**Table 3.**
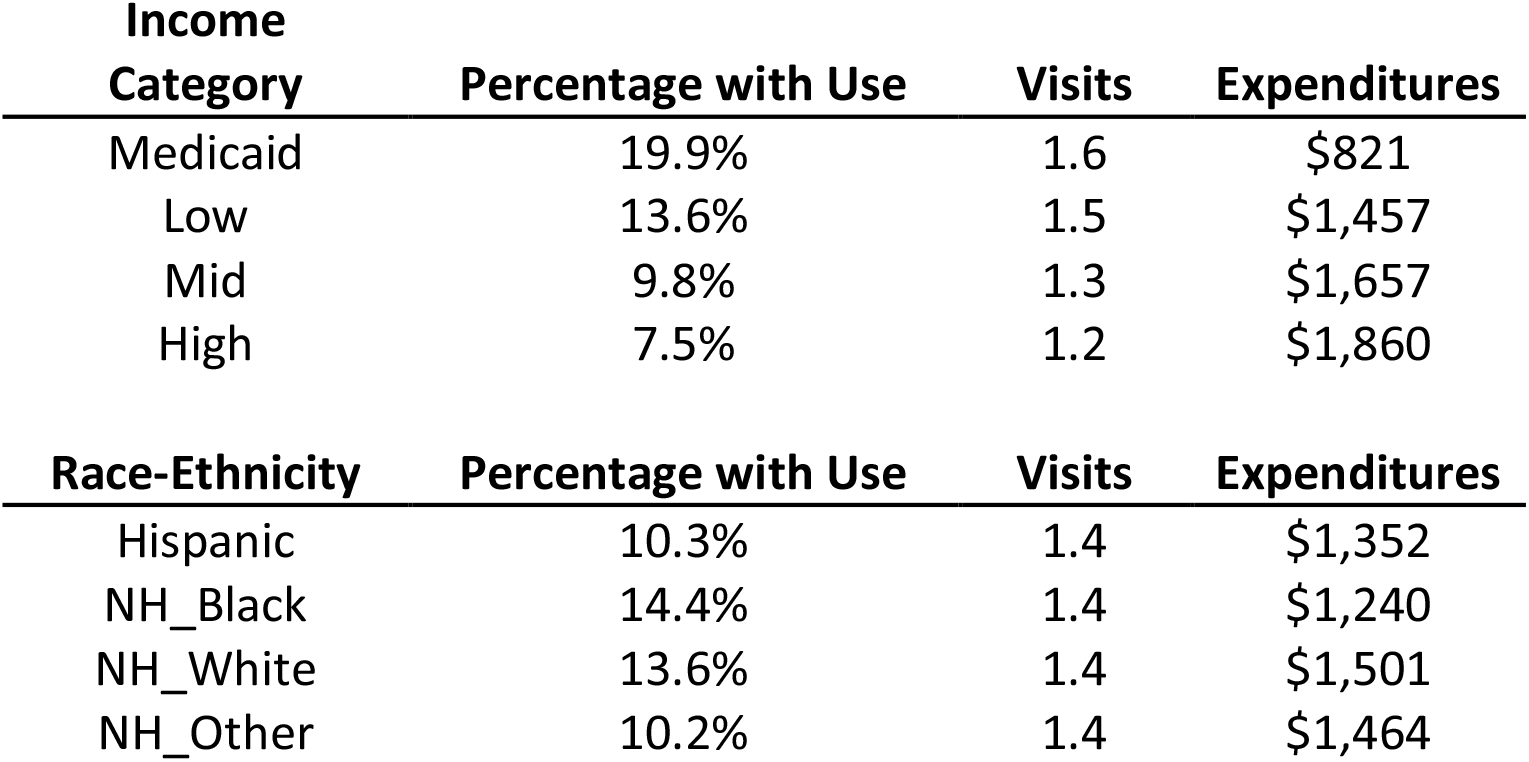
Emergency Room Use and Expenditures by Income Level and Race-Ethnicity.

| **Income Category** | **Percentage with Use** | **Visits** | **Expenditures** |
| --- | --- | --- | --- |
| Medicaid | 19.9% | 1.6 | \$821 |
| Low | 13.6% | 1.5 | \$1,457 |
| Mid | 9.8% | 1.3 | \$1,657 |
| High | 7.5% | 1.2 | \$1,860 |

| **Race-Ethnicity** | **Percentage with Use** | **Visits** | **Expenditures** |
| --- | --- | --- | --- |
| Hispanic | 10.3% | 1.4 | \$1,352 |
| NH_Black | 14.4% | 1.4 | \$1,240 |
| NH_White | 13.6% | 1.4 | \$1,501 |
| NH_Other | 10.2% | 1.4 | \$1,464 |

Overall, 12.0% of the age 0-64 population had emergency room visits and expenditures. Comparing highest and lowest overall emergency room use, those in the Medicaid Income category used 166.6% more than those in the High Income category. Non-Hispanic Blacks had 40.6% more than Others. Among those with emergency room utilization and expenditures, The Medicaid group had the highest number of emergency room visits per person followed by the Low, Middle, and High Income groups with a maximum disparity of 28.1%. The Other race-ethnicity group had the highest number of emergency room visits per person followed by White, Black, and Hispanic; the maximum disparity was 4.5% (Figure 3).

**Figure 3:**
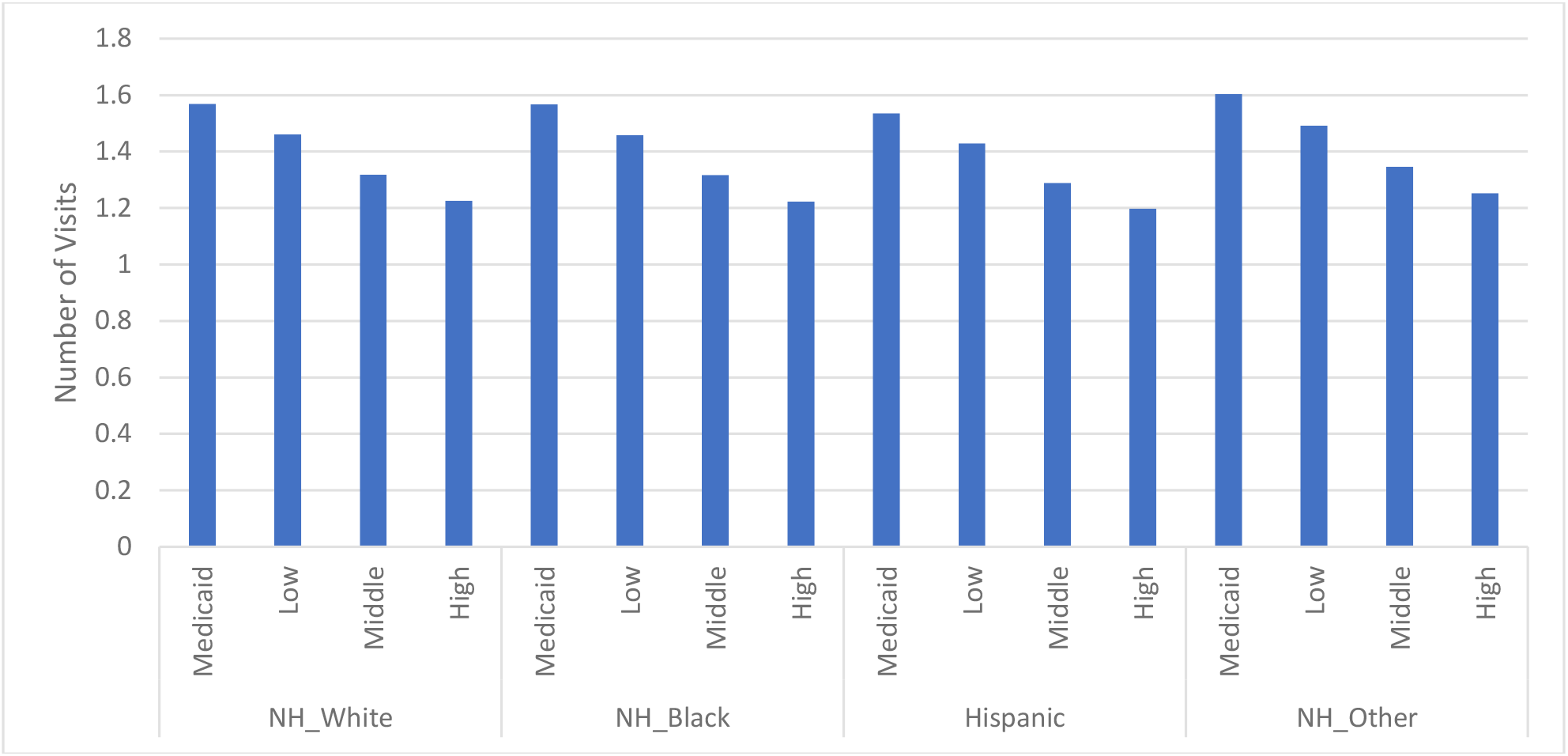
Number of Emergency Room Visits per Person per Year by Income and Race-Ethnicity. (for those with emergency room use and expenditures greater than zero).

For those with any emergency room utilization and expenditures, expenditures were 126.6% higher for the High Income group compared to the Medicaid group. Race-ethnicity differences were quite similar within income groups. The White group had the highest emergency room expenditures per person followed by Other, Hispanic, and Black; the maximum disparity was 21.0% (Figure 4).

**Figure 4:**
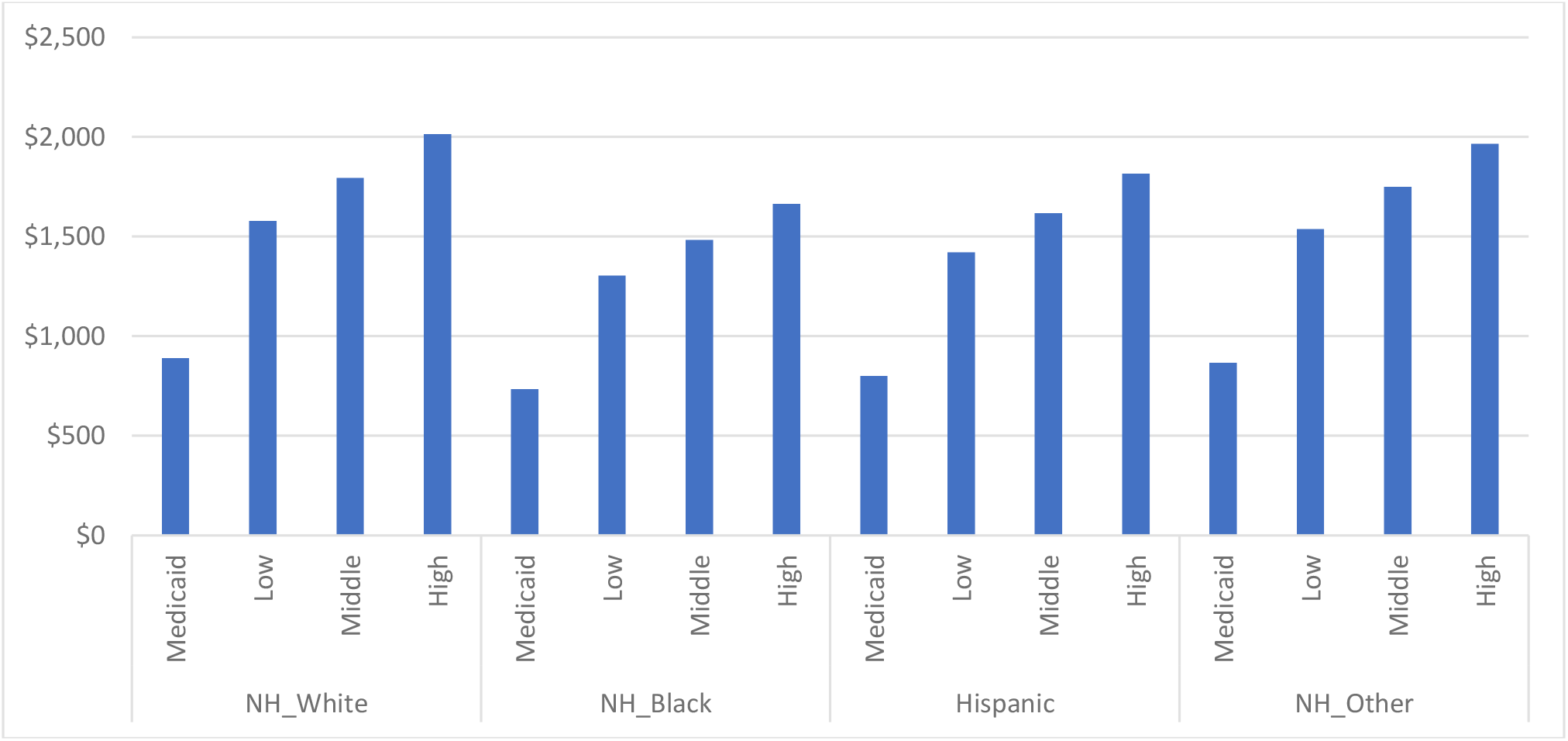
Emergency Room Expenditures per Person per Year by Income and Race-Ethnicity. (for those with emergency room use and expenditures greater than zero).

### Outpatient Visits and Expenditures

Table 4 lists the adjusted percentages of the age 0 to 64 population with any outpatient visits and the average visits and expenditures per person per year for those with outpatient visits and expenditures.

**Table 4.** Outpatient Visits Use and Expenditures by Income Level and Race-Ethnicity.

| **Income Category** | **Percentage with Use** | **Visits** | **Expenditures** |
| --- | --- | --- | --- |
| Medicaid | 73.3% | 9.3 | \$1,839 |
| Low | 59.5% | 5.9 | \$1,587 |
| Mid | 67.6% | 5.3 | \$1,652 |
| High | 74.5% | 6.0 | \$1,915 |

| **Race-Ethnicity** | **Percentage with Use** | **Visits** | **Expenditures** |
| --- | --- | --- | --- |
| Hispanic | 61.7% | 5.8 | \$1,613 |
| NH_Black | 65.6% | 6.1 | \$1,658 |
| NH_White | 78.4% | 7.9 | \$2,123 |
| NH_Other | 68.7% | 6.2 | \$1,626 |

Overall, 72.9% of the age 0-64 population had outpatient visits and expenditures. Comparing highest and lowest overall outpatient use, those in the High Income category used 25.1% more than those in the Low Income category (note that the percentages for the High Income and Medicaid groups were almost identical). Non-Hispanic Whites had 27.0% more use than Hispanics. Among those with outpatient utilization and expenditures, there were 73.1% more outpatient visits for the Medicaid but not the Low Income group compared to the Middle Income group. Whites had the highest number of outpatient visits per person followed by Other, Black, and Hispanic, with a maximum disparity of 35.8% (Figure 5).

**Figure 5.**
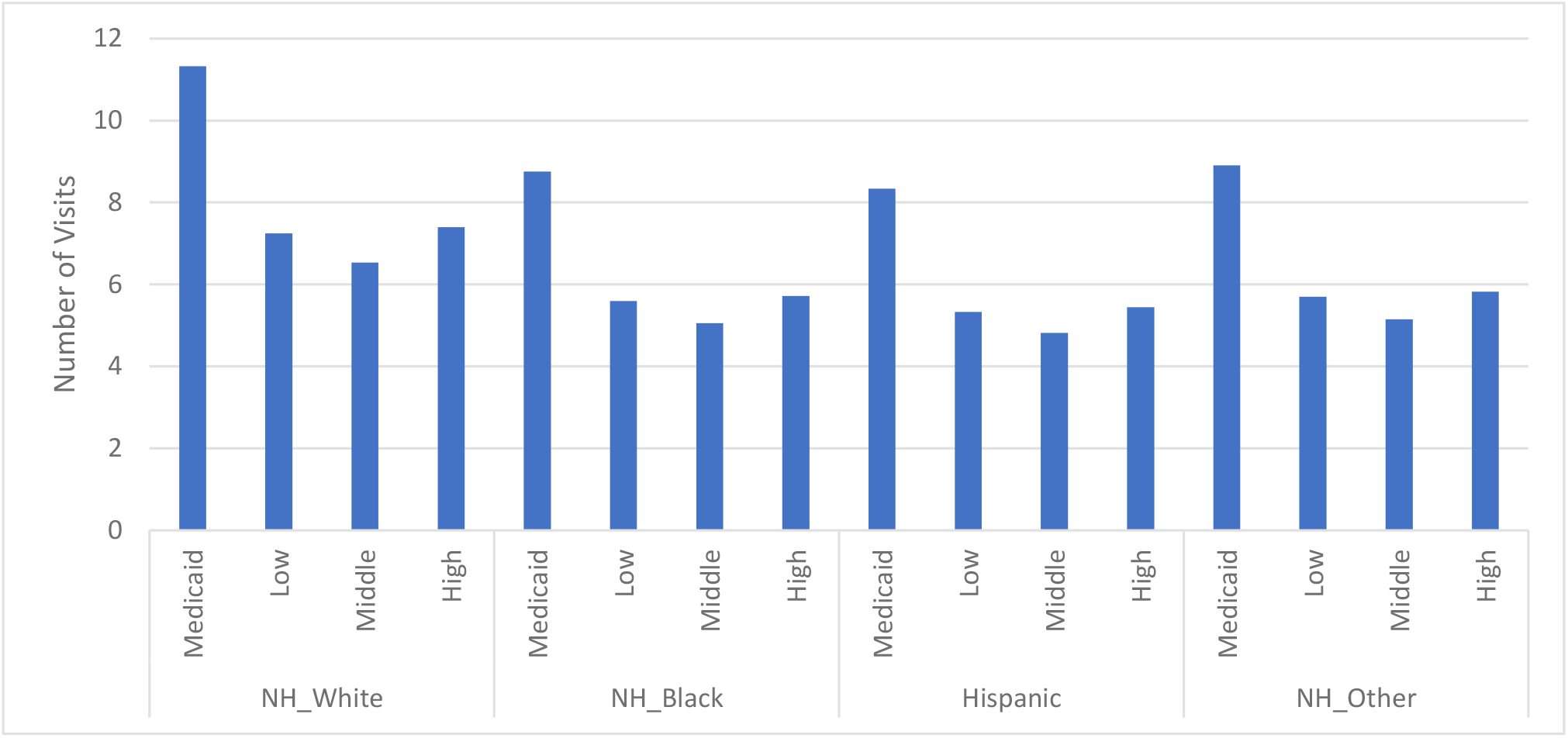
Number of Outpatient Visits per Person by Income and Race-Ethnicity. (for those with outpatient visits and expenditures greater than zero).

Among those with any outpatient utilization and expenditures, expenditures per person were highest in the High Income group but the Medicaid group was only 4.1% less and the maximum disparity was 20.7%. Whites had uniformly higher expenditures with a maximum disparity of 31.6% (Figure 6).

**Figure 6.**
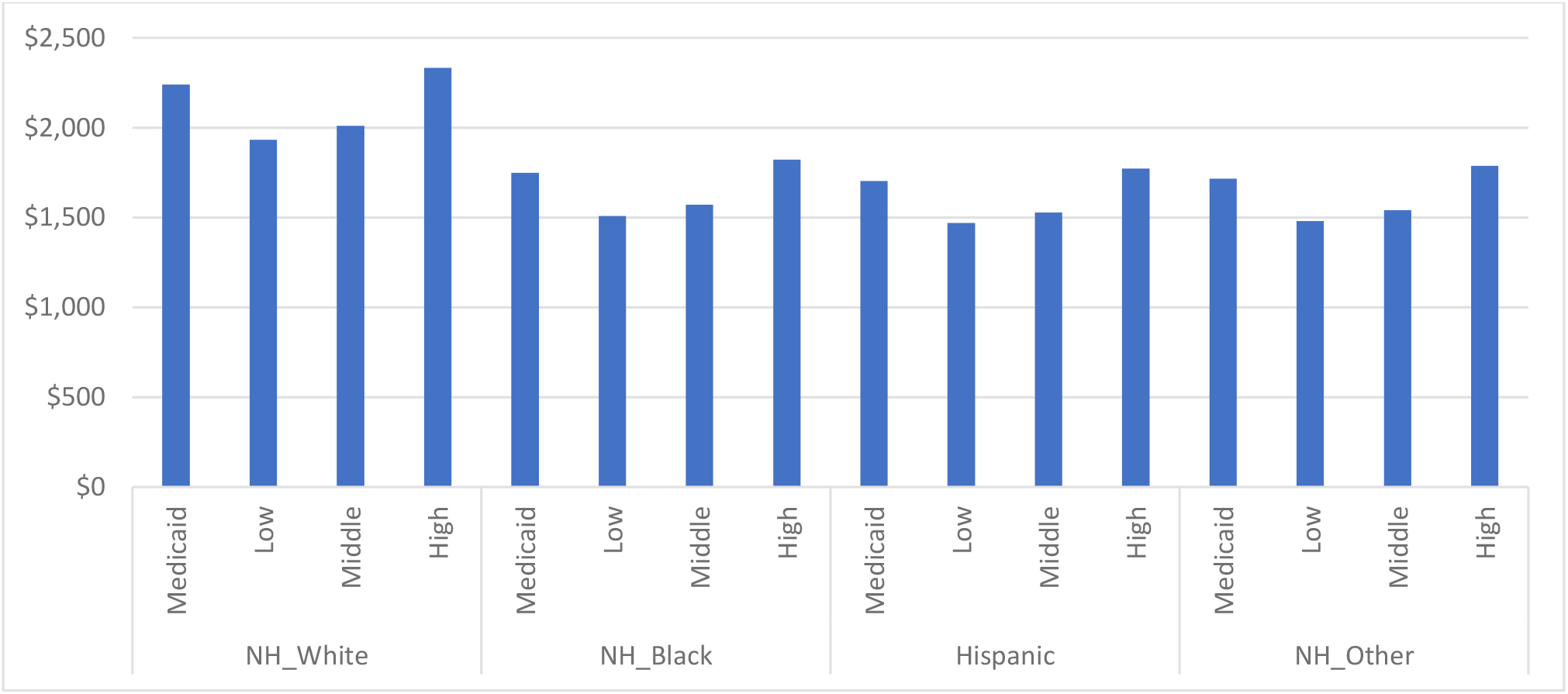
Outpatient Visit Expenditures per Person by Income and Race-Ethnicity. (for those with outpatient visits and expenditures greater than zero).

### Prescription Medication Usage and Expenditures

Table 5 lists the adjusted percentages of the age 0 to 64 population with any prescription medications and the average number of prescriptions and expenditures per person per year for those with prescription usage and expenditures.

**Table 5.** Prescription Medication Use and Expenditures by Income Level and Race-Ethnicity.

| **Income Category** | **Percentage with Use** | **Number of Prescriptions** | **Expenditures** |
| --- | --- | --- | --- |
| Medicaid | 58.0% | 15.9 | \$1,909 |
| Low | 42.2% | 9.1 | \$1,244 |
| Mid | 45.4% | 7.6 | \$1,024 |
| High | 47.8% | 6.1 | \$1,061 |

| **Race-Ethnicity** | **Percentage with Use** | **Number of Prescriptions** | **Expenditures** |
| --- | --- | --- | --- |
| Hispanic | 41.9% | 7.5 | \$1,006 |
| NH_Black | 46.9% | 9.4 | \$1,333 |
| NH_White | 59.8% | 11.0 | \$1,493 |
| NH_Other | 44.7% | 8.7 | \$1,289 |

Overall, 53.7% of the age 0-64 population had prescription medications and expenditures. Comparing highest and lowest overall prescription medication use, those in the Medicaid Income category used 37.5% more than those in the Low Income category. Non-Hispanic Whites had 42.7% more use than Hispanics. Among those with prescription medication utilization and expenditures, the Medicaid group had the highest number of prescriptions per person followed in income order of Low, Middle, and High with a maximum disparity of 160.4%. Whites had the highest number of prescriptions per person followed by Black, Other, and Hispanic with a maximum disparity of 46.2% (Figure 7).

**Figure 7.**
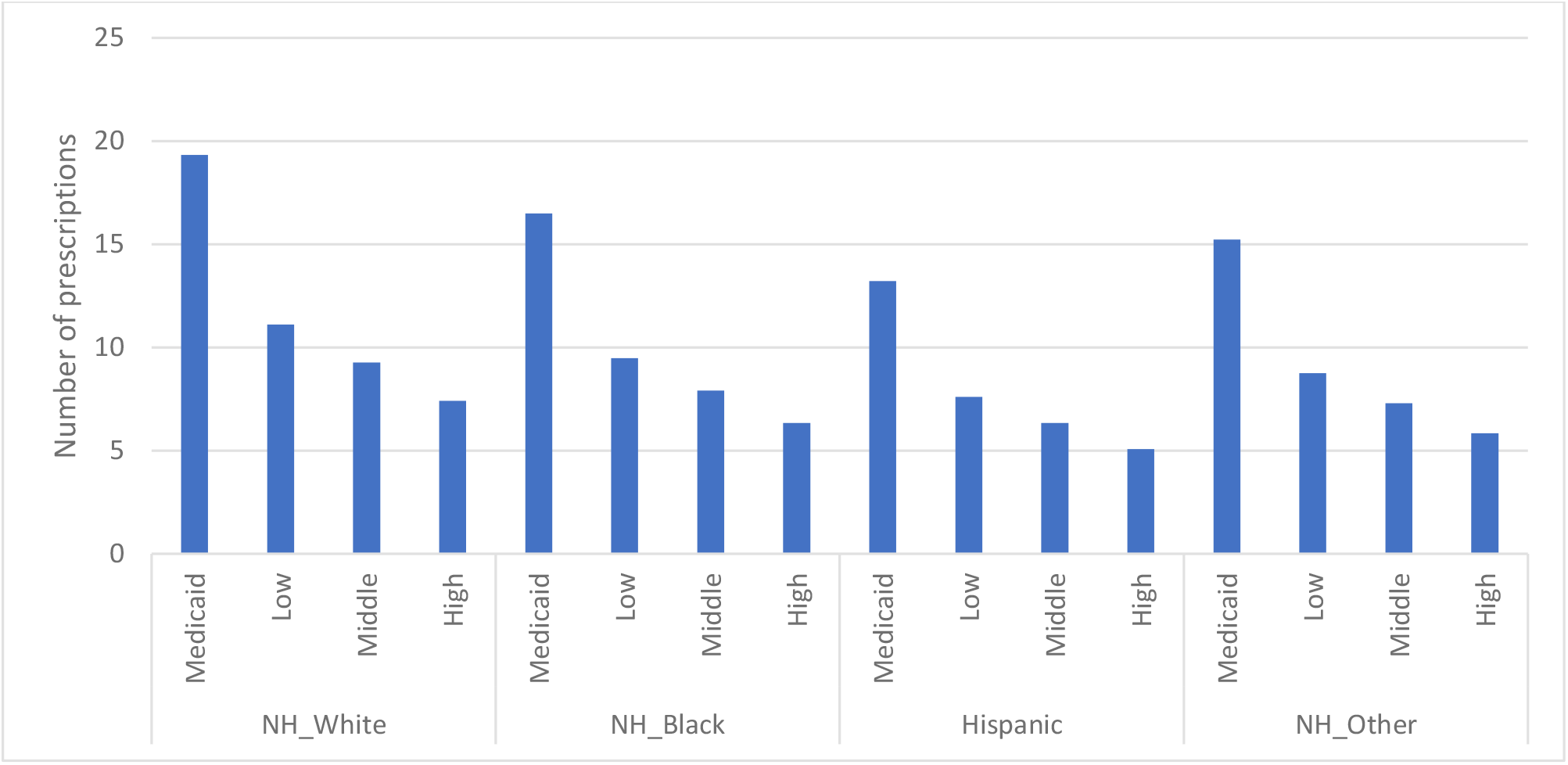
Number of prescriptions per person by income and race-ethnicity. (for those with prescriptions and expenditures greater than zero).

For those with any prescription medication utilization and expenditures, expenditures were 86.4% more in the Medicaid group compared to the Middle Income group. Whites had the highest prescription expenditures per person followed by, Black, Other, and Hispanic. The maximum disparity was 48.4% (Figure 8).

**Figure 8.**
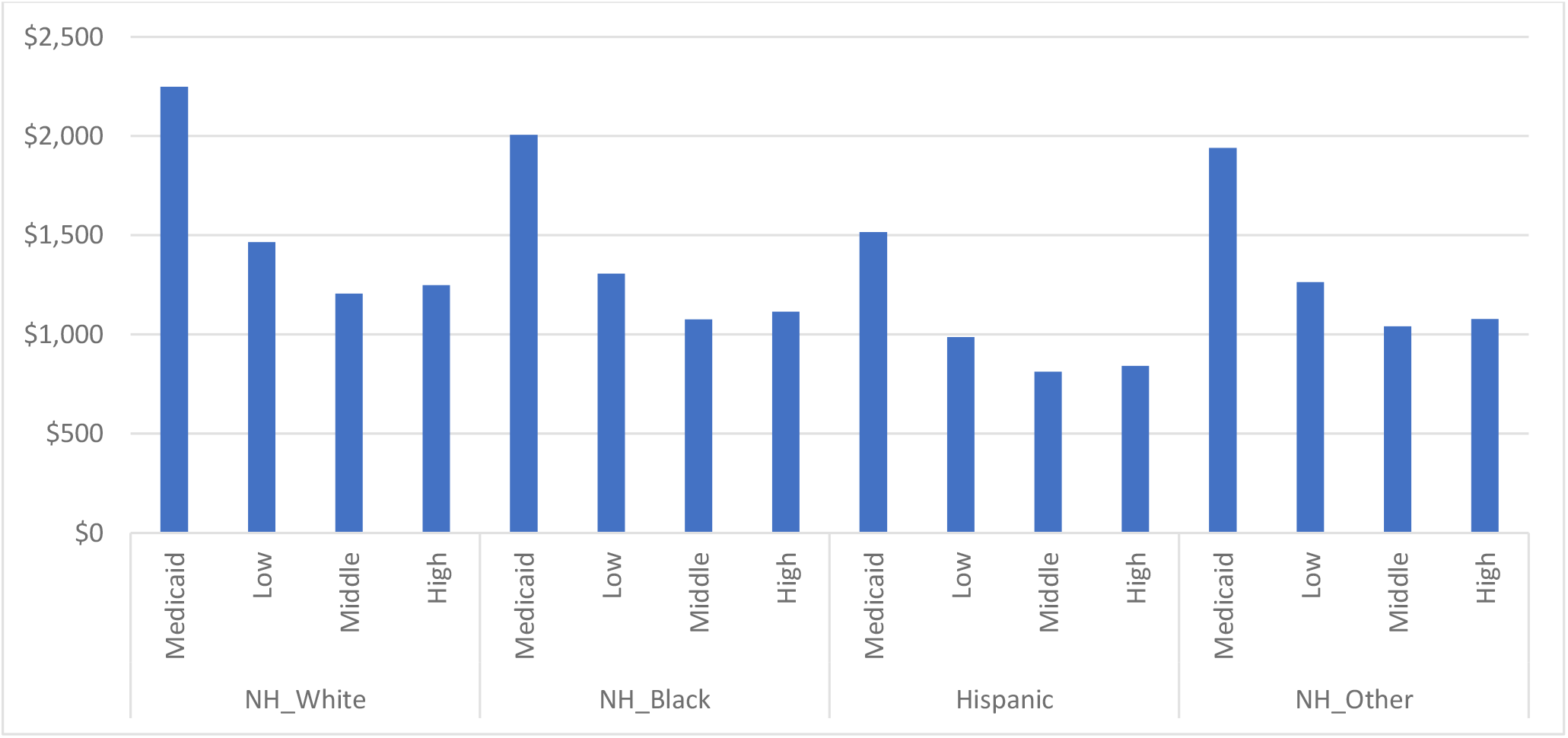
Prescriptions Expenditures per Person by Income and Race-Ethnicity. (for those with prescriptions and expenditures greater than zero).

Table 6 summarizes the results for each of the three measures in each medical service category by income and race-ethnicity.

**Table 6.**
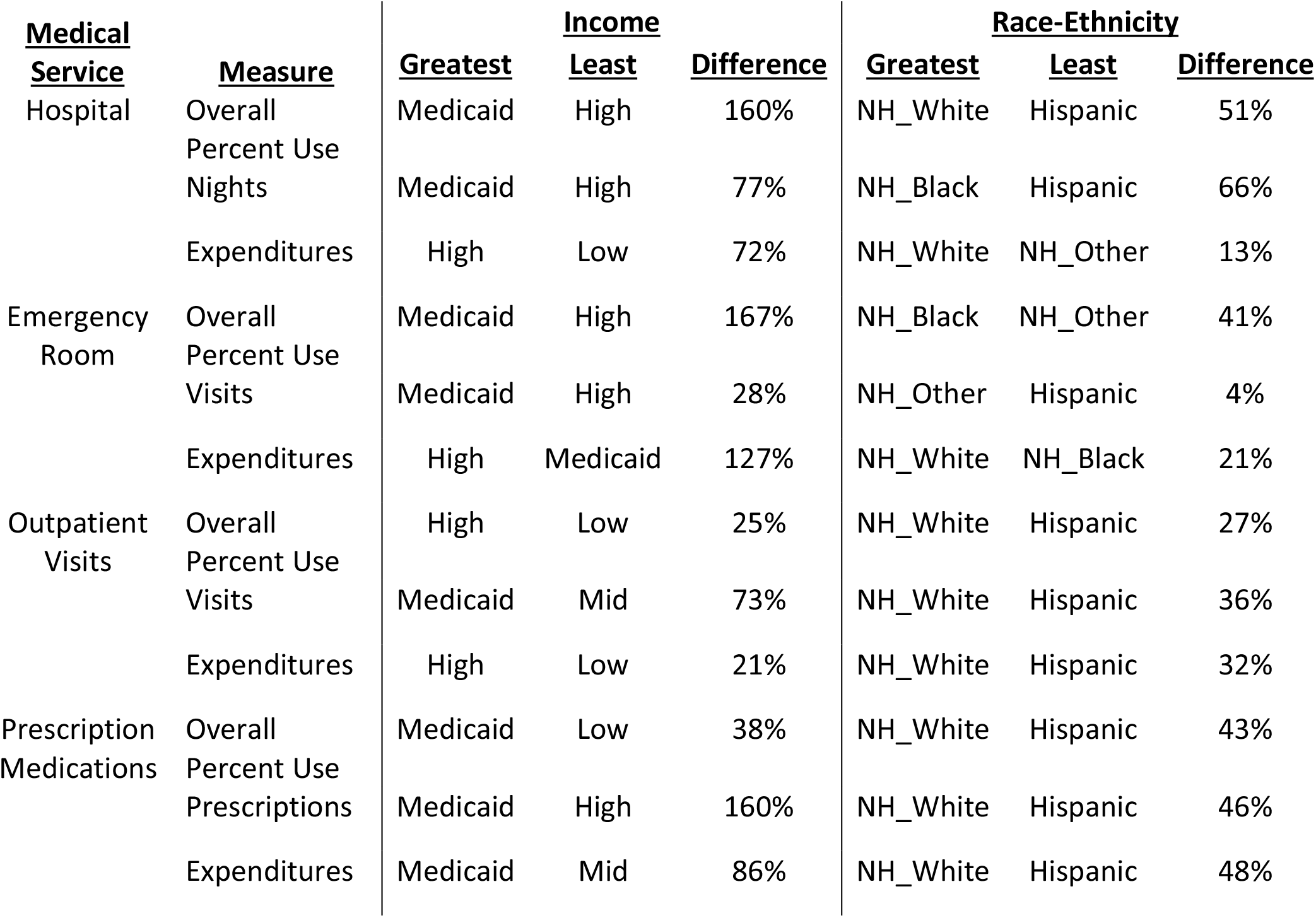
Results Summary.

Of the 12 separate differences listed in Table 6, only 3 of the race-ethnicity differences exceed the corresponding income difference. Two of the three exceptions occurred in the Outpatient Visits category. Moreover, the income differences are generally much greater in magnitude than those for race-ethnicity. Within the income comparisons, those on Medicaid had the greatest utilization in 7 of the 8 comparisons. Conversely, the High Income group had greatest expenditures for 3 of the 4 medical services. Non-Hispanic Whites had the greatest utilization and expenditures for 9 of the 12 measures. Similarly, Hispanics had the least utilization and expenditures for 9 of the 12 measures. The weighted average of differences by income was 56.6% and the corresponding weighted difference by race-ethnicity was 30.7%.

## DISCUSSION

Only 3 of the 12 race-ethnicity differences we examined exceeded the corresponding income difference. Furthermore, income differences were generally much greater than race-ethnicity differences. In general, lower incomes were associated with higher utilization where those on Medicaid had the greatest utilization except for Overall Percent Use of Outpatient Visits. Those with higher incomes had greater expenditures except for prescriptions. Whites had the highest utilization and expenditures in three-fourths of the comparisons and Hispanics were lowest in the same proportion of the comparisons.

Using data from the 2008 NHIS, Dubay and Lebrun found that income disparities in health status, health behaviors, and health care utilization “dwarfed” those based on race-ethnicity.^10^ They also cited previous studies where disparities between socioeconomic groups among adults of the same race-ethnicity were considerably larger than those based on race-ethnicity. More recently, Karmouta and colleagues reported that retinopathy of prematurity (ROP) was more common and severe in Hispanic neonates than in non-Hispanic White neonates.^11^ However, the association between race and ethnicity and ROP was mainly driven by disparities in gestational age, which was mainly explained by neighborhood income level.

Lemstra and colleagues, using data from the Canadian Community Health Survey, found that residents with lower income were responsible for disproportionate usage of hospitals, physicians, and medications.^12^ Yet, contrary to our findings, low income Canadians also had higher medical expenditures. They also noted that most of the disparity in high health care utilization for lower-income residents was associated with higher disease prevalence. Loef, et al, in a study of Dutch adults, reported that low socioeconomic status was associated with increased healthcare expenditures and utilization.^13^ However, the socioeconomic differences largely disappeared after considering health status. They concluded that resources were being spent where they were most needed within the universal Dutch healthcare system. Universal coverage in Canada and the Netherlands undoubtedly contributes to the income related expenditure differences between those countries and the US.

The US has no universal healthcare system for those under age 65. Low Medicaid reimbursement rates and high uninsured rates in the Low income group contribute to lower expenditures for all services except prescription medications.^14,15^ Given the greater expenditures for those with higher incomes in the US and the poorer health of those with low incomes, resources may not be used where they are most needed.^16,17^ Indeed, increased spending on those with high incomes and better health disproportionately benefits them. Although this is perhaps not a design feature of the US “system,” it reflects the current power dynamic where the affluent have much more influence on legislative and corporate decisions.

Education is often touted to achieve socioeconomic equity. Yet the pandemic taught us that many essential jobs do not require higher education—home delivery, day care, nursing home assistants, and slaughterhouse workers to name a few. Thus, more education should not be considered a panacea for income inequality.

We suggest that income in this study is a marker for a range of social factors that affect health. The US Preventive Services Task Force found that the most important social risk factors are food insecurity, housing instability, and transportation, each of which eventually involves income.^18^ Notably, Medicaid and Medicare are taking steps to address socioeconomic determinants of health.^19,20^ There are challenges and pitfalls associated with prioritizing various social determinants of health. Causal mechanisms of the relationships between health and income disparities, other social factors, their interactions, and effect sizes need further investigation.

Organized medicine should also play a role in reducing disparities. American Medical Association (AMA) President, Gerald E. Harmon, MD, advocated for greater emphasis on social determinants of health to stem the decline in life expectancy in the US.^21^ While we laud his stance, it remains to be seen whether the AMA and physicians will back increased social spending that may limit increases in medical spending. In that regard, Adler and colleagues wrote, “Expenditures on clinical care have an opportunity cost, and the amount of money devoted to health care delivery makes it difficult to provide sufficient support for other kinds of investment that would have greater health benefits.”^22^

Improving health in the US will require a multifaceted approach; we suggest beginning with changes in healthcare. The medical community needs to question the effectiveness of the current healthcare “system” for those under age 65 and refocus the current research on socioeconomic factors to create a cost-effective system for all. Although specific recommendations on changes are beyond the scope of this article, the guiding principle must be to use resources where they are most needed. For example, Ritchie and Leff described the value of home-based care.^23^ Once we have an efficient healthcare system for those under 65, then freed up resources can be used to attack problems such as food insecurity, housing instability, transportation, and child care. Typically, the medical community rapidly and aggressively addresses a known factor that results in morbidity and death. Why is income disparity different?

## Limitations

Since most MEPS data come from self-reports, differential reporting and selective non-response between the various income and race-ethnicity groups is possible. It is also important to recognize that the category NH Other is a mixture of mostly Asians and Native Americans and Alaskans, groups with considerably different medical care use and expenditures.^24^ We did not have sufficient data to make stable estimates for each sub-group. In addition, MEPS is unable to sample high-cost hospital and physician expenditures that occur just before the sampled person dies or is placed in a nursing home resulting in underestimates. This consideration should be less applicable in the 0 to 64 age group. Another potential limitation is that some of the respondents could be disabled and have Medicare coverage despite being less than 65 years of age. Finally, we recognize that it is impossible to design a study or statistically adjust away all confounding leaving residual confounding between income and race-ethnicity.

These data include no information on the quality or optimal amounts of care. Optimal amounts of care surely differ by income as well as race-ethnicity. The measure of inequality, the percent difference between the group with the lowest utilization or expenditures and the group with the highest utilization or expenditures, uses only two of the four values in any income or race-ethnicity subgroup. Measures such as slope can use all four values but are still subject to outliers. We submit that our method is straightforward and consistent with our goal in this exploratory analysis.

## CONCLUSIONS

These results indicate that income inequalities are more strongly associated with medical care utilization and expenditures than race-ethnicity among those aged 0-64. However, the results do not include adjustment for health. The next step is to adjust for the health status and comorbidities of the individuals as those factors are more germane to health outcomes and expenditures than race-ethnicity and income. Although more research should focus on income related health disparities in the United States, it is time to recognize that sound health policy must include reducing socioeconomic inequalities, especially those related to income.

## Data Availability

Data are in the public domain and may be downloaded from: https://meps.ipums.org/meps/

## Notes

### Competing Interest Statement

The authors have declared no competing interest.

### Funding Statement

This study did not receive any funding.

### Author Declarations

IPUMS Health Surveys MEPS https://meps.ipums.org/meps/

